# Life Beyond the Forensic Unit: A Systematic Review and Meta-analysis of Patient Reoffending, Hospital Readmission, and Mortality Rates Following Discharge to the Community

**DOI:** 10.64898/2026.05.27.26354062

**Authors:** James McLauchlan, Carey Marr, Richard Kemp, Kimberlie Dean

## Abstract

**Background:** Forensic patients often have complex and costly healthcare needs and risks, even following discharge from secure care. However, little is known about these patients’ health and justice outcomes after community reintegration.

**Aims:** To estimate the incidence of key post-discharge outcomes among community-discharged forensic patients, including any reoffending, violent reoffending, reconvictions, readmissions, all-cause mortality, and suicide.

**Method:** We systematically searched PsycINFO, Embase, CINAHL, Medline, PubMed, and ProQuest Dissertations from database inception to May 2025 (PROSPERO CRD42024529265). Random-effect meta-analyses were used to generate pooled incidence estimates, with heterogeneity quantified using prediction intervals.

**Results:** A total of 49 studies met inclusion criteria (*n* = 18,871) and contributed to the meta-analyses. The pooled incidence rate per 100,000 person-years was: any reoffending 3,889 (95% CI 2,055–7,359; 95% PI 290–52,136); violent reoffending 1,851 (95% CI 1,229–2,789; 95% PI 201–17,068); reconvictions 3,291 (95% CI 2,591–4,179; 95% PI 950–11,394); readmissions 7,945 (95% CI 5,507–11,463; 95% PI 1,225–51,548); all-cause mortality 1,789 (95% CI 1,341–2,388; 95% PI 673–4,756); and suicide 407 (95% CI 319–519; 95% PI 225–735).

**Conclusion:** Overall, the reoffending rate for forensic patients discharged to the community waslower than that reported for other cohorts of people charged with general and violent offences. However, despite typically receiving long admission periods, discharged forensic patients continue to experience high rates of readmission, all-cause mortality, and suicide relative to other psychiatric patient groups in the community. Together, our findings highlight a need for enhanced post-discharge suicide support for forensic patients living in the community to better facilitate successful, long-term reintegration.

---

Forensic patients discharged into the community pose a complex challenge for both public mental health and criminal justice systems. In countries with established forensic mental health treatment pathways, including the United Kingdom (UK) and Australia, people with serious mental illness, particularly schizophrenia, account for 51.9% to 83.9% of forensic patient cohorts^1,2^. Further adding to the complexity is that most individuals admitted to secure care facilities have other comorbidities including substance use problems, cognitive impairment and personality disorders^2^, all of which contribute to an increased risk of adverse post-discharge outcomes, including violent offending ^3^. Therefore, any meaningful evaluation of forensic mental health care must assess not only its effectiveness in improving health outcomes, but also in reducing future offending and supporting public safety.

## Patient Complexity

Forensic patients commonly experience difficulty adjusting to community life after treatment. Fazel et al.^4^ conducted a 15-year follow-up study of 6,520 Swedish forensic patients to examine their trajectory after discharge from care. By the end of the follow-up period, over two-thirds (69%) were rehospitalised at least once, two-fifths had been charged with a new violent offence, and a fifth (22%) had died by suicide. Forensic patients are more likely to have severe and treatment-resistant psychosis, making intervention particularly challenging. For example, in Ireland, a quarter of patients in the national forensic mental health service were found to meet criteria for a treatment-resistant psychosis diagnosis^5^. In addition to psychotic symptoms, they commonly present with comorbid conditions that add to the treatment complexity, including substance use problems, personality dysfunctions, and cognitive disabilities^2^. Forensic patients are also at risk from several social adversity in the community, including socioeconomic disadvantage and reduced access to psychiatric care when not mandated, making them more susceptible to mental health deterioration^6–8^. As such, forensic patients require an intensive and coordinated multidisciplinary treatment, with a high level of post-discharge monitoring and support.

## Offending Risk

The increased offending risk amongst people with severe mental illness arises from a complex interplay between psychotic symptoms that impair judgement and precipitate affect disturbances, and co-occurring social challenges^9,10^. Fazel et al.^11^ conducted a meta-analysis of 20 studies examining the association between schizophrenia and violence. People with psychotic symptoms was almost 20 times more likely to commit a serious violent crime than the general population. This association is not limited to risk of criminal justice system contact for violent offending, with the risk of any offending for people with psychosis found to be twice that of people without psychosis^12^. Collectively, these findings regarding health and justice risks highlight the need for forensic patients to have adequate support and monitoring post-discharge to the community, and for the ability of forensic mental health services and their partners to meet such need to be rigorously and regularly evaluated.

## Forensic Mental Health Services

Forensic mental health care services occupy a difficult but important role in society. These services are responsible for providing treatment to patients with serious mental illness who are in contact with the criminal justice system. Alongside the challenges of rehabilitation for patients presenting with significant mental health complexity, forensic services must also balance the rights of patients not to be detained longer than necessary and their obligation to protect the patients and the public from foreseeable harm. Unsurprisingly, such services have drawn criticism, including keeping patients longer than warranted, on the one hand, and failing to protect the public, on the other, as well as for being costly to operate ^13^. The annual cost per patient for forensic psychiatric care in the UK, for example, was estimated to be approximately £175,000 in 2008 ^14^. More recently, in Australia, the average cost of caring for a single forensic patient in 2024 was suggested to be roughly $537,000 AUD per year ^15^. The substantial costs of forensic mental health care necessitate careful evaluation of post-discharge health and justice outcomes to ensure these costly interventions are leading to better patient outcomes and improved public safety.

## Forensic Patient Outcome Evaluations

Several limitations currently exist in the extant literature on outcomes for people discharged into the community from forensic mental health services. First, despite the clear need for an integrated perspective considering both justice and health outcomes, most research has focused predominately on reoffending rates, with far fewer studies exploring other indicators, including suicide rates and post-discharge re-admission^16^. The second issue concerns the highly inconsistent findings across studies, even within similar jurisdictions, with reported reoffending rates as high as 21% ^17^ and as low as 6.3%.^2^. Another problem is that most outcome evaluation studies are underpowered, with many cohorts containing fewer than 100 ^18,19^ ^20–22^ and less than a handful of studies have samples over 1,000 ^4,23,24^. The fourth issue is the wide variability in study designs. Crucially, there is little consistency in follow-up reporting periods, ranging from 12 months ^2^, to 4 years ^25^, and up to 20 years ^26^. Collectively, these gaps provide an incomplete picture of the challenges facing patients after discharge and the effectiveness of forensic mental health services, limiting the generalisability of findings beyond individual study settings.

Hitherto, only one meta-analysis has synthesised the outcomes of patients discharged from secure psychiatric facilities. Over a decade ago, Fazel et al.^16^ synthesised 35 studies on discharged forensic patients across 10 countries (*n =* 12,056). They reported a crude reoffending rate of 4,484 per 100,00 person-years, alongside a readmission rate of 7,208 per 100,000 person-years. For health outcomes, the all-cause mortality rate was 1,538 per 100,000 person-years, while the suicide rate of 325 per 100,000 person-years. However, as the authors noted, several studies in their -analysis did not provide sufficient detail about patient discharge destinations and thus estimates of outcomes post-discharge to the community could not be calculated. The pooled estimates were derived from samples that likely included people who laterally discharged to prisons or other secure inpatient settings and never ceased supervision. Including such people in the analysis could unintentionally inflate intervention effectiveness and/or underestimate rates of adverse outcomes.

## Aims

The present meta-analysis aims to provide an up-to-date and comprehensive synthesis of key health and justices outcomes among forensic patients discharged *to the community*. To this end, we calculated incidence estimates for any reoffending, violent reoffending, reconvictions, readmissions, all-cause mortality, and suicide to provide a comprehensive understanding of community trajectories in this population.

## Methods

This systematic review and meta-analyses were pre-registered on PROSPERO on March 26th, 2024 (CRD42024529265). The meta-analysis was conducted and reported following the recommendations outlined in Preferred Reporting Items for Systematic Reviews and Meta-analyses (PRISMA; Moher et al., 2009) and the Meta-analysis of Observational Studies in Epidemiology (MOOSE; Stroup et al., 2000; see Appendix B for full checklist) guidelines.

### Search Strategies

The systematic review searched the following databases: PsycINFO (Ovid), EMBASE (Ovid), CINAHL (EBSCO), MEDLINE (Web of Science), PubMed, and ProQuest Dissertations & Theses Global (Web of Science). The first database search was conducted on May 6^th^, 2024, with an updated scan performed on May 16^th^, 2025, to identify recently published studies. No restriction was applied regarding language, publication date, or reference type. Databases were searched using keywords related to the target sample ("forensic patients" or "patients" or "mentally ill offend*" or "mental disord*" or "hosp* patients", psych* disord*), institution ("forensic hosp*" or "forensic ment*" or "low secur*" or "medium secur*" or "high secur*" or "or "psych* hosp*" or "forensic rehab*"), and the key health and justice outcomes ("Mort" or "Rehosp*" or "Readm*" or "Death" or "Reconvict*" or "Reoffend*" or "recidi*" or "rearrest" or "repeat offend"). This search strategy was developed in collaboration with the University of New South Wales Librarian team to maximise coverage while maintaining efficiency. All citations identified using these search terms were exported to Covidence, software for support systematic reviews and meta-analyses for screening. Details of the screening process are presented in the PRISMA flowchart (Figure 1).

**Figure 1.**
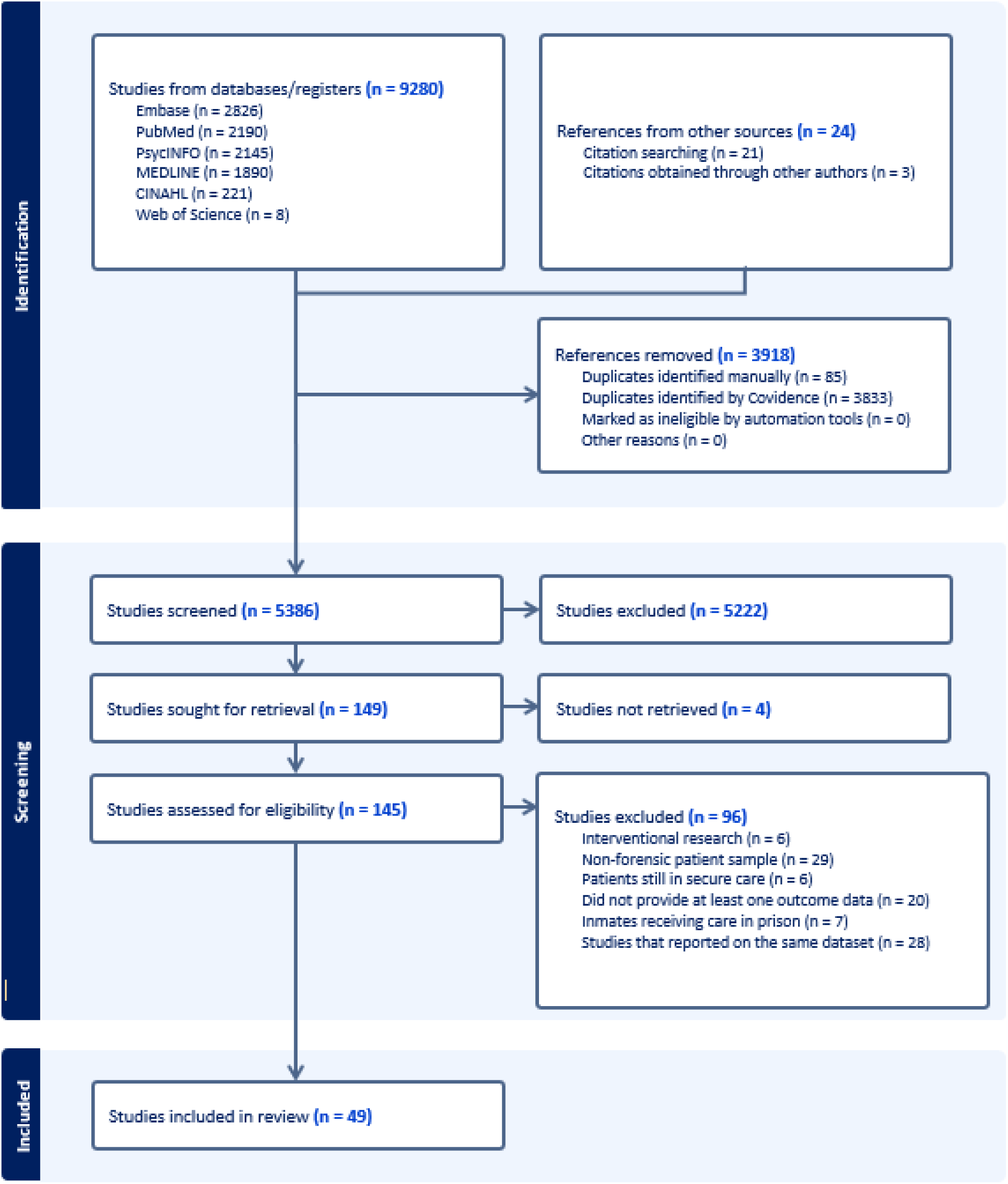
PRISMA diagram. The flowchart summarises total number of studies initially identified, screened, met eligibility criteria, and included for the meta-analysis.

### Inclusion and Exclusion Criteria

To be included in the systematic review, studies had to meet the following criteria: (a) Reported data on patients discharged into the community from any secure forensic psychiatric hospital, including high-, medium-, or low-security facilities, either in published peer-reviewed journals, government reports, or theses; (b) reported on the count or incidence rates of at least one adverse outcome, including reoffending, reconvictions, readmission, all-cause mortality, and suicide in person-years data or the mean/median follow-up duration. To ensure the estimates meaningfully reflect the adverse outcomes patients experience after discharge to the community, we only included discharged cohorts with confirmed community access. When studies reported outcomes separately for a full discharged cohort and a subgroup with verified community access, we extracted data only from the latter.

The exclusion criteria for the review were as follows: (a) studies of patients discharged from other non-secure psychiatric or correctional facilities; (b) studies that reported on the same outcomes using an overlapping dataset; and (c) studies on forensic patients that did not provide at least one community-based discharge outcome; (d) studies of people who have offended and are receiving mental health treatment in prison only; and (e) intervention studies.

Publications written in a language other than English were included in the systematic review to ensure representativeness. A standardised procedure was followed when screening non-English articles. First, initial translations were conducted using Microsoft Translate, and key information related to study eligibility and outcome variables was extracted. This information was then confirmed with the study authors via email. When study authors were unavailable, a postgraduate student in the University of New South Wales School of Psychology who was a native speaker of the relevant language was identified to review the translated material to ensure accuracy and identify any errors.

### Screening and Data Extraction

The first (J.M.) and second authors (C.M.) performed the systematic review screening. After removing duplicates, the two investigators independently screened titles and abstracts against the eligibility criteria. A full-text review was then conducted for the studies that passed the first stage screening to confirm eligibility. Any discrepancies with the selection were resolved through discussion, and where an agreement could not be reached, the project supervisor (K.D) made the final decision. Study authors were contacted if there was missing data, inadequate demographic descriptions, or when full-text copies were unavailable. On these occasions, each study author was contacted twice, with at least two weeks between emails, and given until the end of data extraction, beginning on June 2^nd^ and finished on June 30^th^, 2025, to respond. The reference sections of eligible papers were also reviewed to identify potentially relevant papers otherwise missed.

### Data Extraction

The first author, J.M., performed the data extraction. Any ambiguity with the reported data in studies was resolved through discussion with the project supervisors, C. M, R.K, and K.D. Sociodemographic characteristics and study details were extracted for moderator analyses where information was available. The list of variables extracted from each study included: country of origin, institution, admission periods, publication year, publication type (e.g., peer-reviewed journal), security level (i.e., high, medium, low security), sample size, mean age, gender, primary diagnosis (e.g., schizophrenia), mean or median follow-up duration, total year spent at risk, and study outcomes reported (e.g., violent reoffending).

### Outcome Measures

To ensure consistency when extracting data, all six post-discharge outcomes were defined using explicit criteria. Reoffending was categorised as *Violent* if it met the United Nation’s international Classification of Crime for Statistical Purposes (ICCS) ^29^ definition of violence. These include acts leading to death, intent to kill, acts causing harm, sexual acts, and property crimes with violence. *Any reoffending* was defined as any type of offending, violent or non-violent, occurring in the community, including that leading to arrests or charges, not yet resulted in formal convictions. *Violent reoffending* included the following specific offences: homicide, assault, abduction, robbery, harassment, threats, stalking, and all contact sex offences. While there was an intention to separately report rates of *non-violent reoffending,* there were insufficient studies available (*n* = 6). *Reconviction* included all offences committed in the community that resulted in a formal guilty or a not criminally responsible due to reasons of mental illness verdict, whatever the disposal. Offences committed whilst not in the community were excluded. *Readmissions* were classified as any voluntary or involuntary psychiatric hospitalisation following discharge into the community, irrespective of whether it was to the discharging hospital or other psychiatric facility. *All-cause mortality* included any deaths recorded post-discharge to the community with *suicide* based on reported rates of confirmed cases. All outcomes were treated as binary variables.

### Methodological Quality and Risk of Bias

Each study included for data extraction was assessed for methodological quality and risk of bias using the Joanna Briggs Institute (JBI) Critical Appraisal Checklist for Studies Reporting Prevalence Data ^30^. The JBI checklist contains nine items that help identify potential sources of bias present in study recruitment, design, or analysis. Each item is rated using a binary format (i.e., yes or no) with the option of “unclear” or “not applicable” where needed. The total score for each study can range from 0 to 9, with higher scores indicating a lower likelihood of methodological concerns and risk of bias. Methodological quality was categorised as low (≤3), moderate (4–6), or high (≥7) depending on the number of checklist items met.

### Establishing Independent Samples when Pooling Estimates

A key assumption for meta-analysis is the independence of observations (i.e., individual studies). However, given that one of the study’s aims was to establish an overall incidence rate across several key outcomes, individual studies addressing different outcomes were included in more than one analytic model, but such analyses were undertaken separately, preserving the independence of observations within each outcome. Mistakenly drawing on studies reporting the same outcome with overlapping datasets does risk violating the independence assumption and thus it was necessary to establish a review procedure for identifying distinct studies and managing overlapping datasets.

We employed a structured strategy for verifying independent samples and determining which to include in the review. During data extraction, we routinely collected study details that could indicate overlap, such as the institution, authors’ affiliations, and the admissions period. Studies with similar characteristics were then compared in detail to assess whether they represented the same cohort. In cases of overlap, study selection was based on (a) having a larger sample size, or (b) having more detailed descriptive data about the sample where the sample size was identical. Study authors were contacted for clarification in cases where information was insufficient.

### Effect Size Calculation

Effect sizes based on total person-years were extracted for each study when reported. Person-years was selected as the metric, rather than proportion offending over a specific time period because incidence rates better capture the magnitude of the time patients spend at risk and enable direct comparison between samples or studies ^31^. When person-year outcomes were unavailable and the study was published within the past 15 years and thus more likely to be derived from accessible data, authors were contacted directly. In the absence of a response or the study was more than 15 years old, crude person-years at risk were estimated by multiplying the number of events (e.g., readmissions) by the mean follow-up duration. The mean-based method has been commonly used in prior meta-analyses when the total person-years data were unavailable ^32,33^. The median follow-up duration was used only when the mean was not reported because reliance on the former approach tends to significantly under-estimate total person-years ^34^.

### Statistical Analysis

An incidence rate was calculated for the following post-discharge to the community outcomes: any reoffending, violent reoffending, reconviction, readmission, all-cause mortality, and suicide rate. Each pooled effect size was calculated using a random-effects model to account for expected variability across studies and to increase the generalisability of estimates beyond the cohort studied ^35^. For outcomes with sufficient samples (*n* > 10), we ran additional post-hoc random-effects models restricted to studies published after 2013 in order to enable comparisons with a previous review of outcomes not limited to those confirmed to have occurred after discharge to the community ^16^. See Supplementary material (Appendix A) for more details of the post-hoc analyses. All analyses used the inverse variance method with a restricted maximum-likelihood estimator for between-study variance (τ²). A log transformation and normal approximation confidence interval were calculated for individual study estimates. Given that the I^2^ statistic and Q do not convey the magnitude of heterogeneity or the likely range of true effects, we presented the 95% prediction intervals, following Borenstein’s ^36^ approach. Funnel plots are routinely used to assess potential publication bias and identify asymmetry related to statistical significance (i.e., null vs. positive results) in meta-analyses. However, because this review focuses on synthesising incidence rates, which do not have a natural zero, funnel plots would be inherently asymmetric and, therefore, not a useful measure for assessing bias in this context ^37^. For this reason, funnel plots were not used. Effect sizes (i.e., incidence rates) are reported as events per 100,000 person-years with 95% confidence intervals. All analyses were conducted in R Studio using the Metafor ^38^ package.

Subgroup analyses were conducted to examine potential sources of between-study heterogeneity. The Cochrane Handbook for Systematic Reviews of Interventions ^39^ recommends including at least 10 observations per moderator to avoid overfitting in meta-regression models while ensuring meaningful differences can be detected. Accordingly, subgroup analyses were only performed for outcomes with more than 10 studies, which included violent reoffending, any reoffending, reconvictions, and readmissions. Following this guideline, we selected continuous moderators that are commonly associated with differences in outcome estimates across studies. For outcomes with 10 to 19 studies, only the mean follow-up duration was used as a moderator in the mixed-effect meta-regression model. For those with 20 or more studies, both the mean follow-up duration and proportion of patients with schizophrenia spectrum disorders (SSD) were included as moderators. A significance level of *p* < .01 was adopted to reduce the likelihood of Type I error due to multiple comparisons.

## Results

As displayed in Figure 1, our database searches identified 9,305 citations before duplicates were removed. Of those, 149 papers were eligible for full-text review after completing the title and abstract screening (Cohen’s kappa = 0.61). A total of 49 papers remained after removing ineligible studies and overlapping patient samples (Cohen’s kappa = 0.60, Table 1). The 49 studies included 18,871 patients across 14 countries, contributing to approximately 197,114 total person-years of observation. The studies were primarily conducted in Western countries, with a majority from the United Kingdom (*n* = 17), followed by the United States and Canada (*n* = 5 each). Other countries included Sweden (*n* = 4), Italy and Japan (*n* = 3 each), Australia and Finland (*n* = 2 each), as well as Germany, France, Denmark, and Israel (*n* = 1 each). Sample sizes varied considerably, ranging from 33 to 6,505 patients, and post-discharge follow-up duration ranged between 1 to 19.2 years (*M* = 6.34 years). Most cohorts were predominantly male, with proportions ranging from 65.9 to 95.4%, except for two women-only studies. Mean patient age ranged from 29.3 years to 47.3 years, with an overall mean age of 37.51 years. The number of studies reporting on each outcome was as follows: any reoffending (*n =* 13), violent reoffending (*n* = 28), reconviction (*n* = 29), readmissions (*n* = 24), all-cause mortality (*n* = 10), and suicide (*n* = 8).

**Table 1.** Background characteristics of patient outcome studies included in the meta-analysis (*n* = 49).

| Author, year | Country | Institution/Data registry | *n* | % Male | *M* age | Sample with community access | % SSD | Reoffending data source | Readmission data source | Mortality data source |
| --- | --- | --- | --- | --- | --- | --- | --- | --- | --- | --- |
| Silver et al., 1989 | United States of America | Clifton T. Perkins Hospital* | 127 | 100 | 31 | 127 | 70.9% | Case file reviews | n/a | n/a |
| Bailey & Macculloch, 1992 | United Kingdom | Park Lane Advanced Psychiatric Unit | 106 | 100% | Unavailable | 106 | Not reported | Home Office Offender Index | n/a | n/a |
| Tellefsen et al., 1992 | United Kingdom | Clifton T. Perkins Hospital* | 60 | Unavailable | 33 | 60 | 61.7% | n/a | Case file reviews | n/a |
| Cope & Ward, 1993 | United Kingdom | Reaside Clinic. West Midlands | 51 | 74.5% | 39 | 35 | Not reported | Home Office Offender Index | Case file reviews | n/a |
| Russo, 1994 | Italy | Barcelona Maximum Security Hospital, Messina | 91 | 100% | 40.3 | 91 | 47.3% | Case file reviews | n/a | Case file reviews |
| Baxter et al., 1999 | United Kingdom | Bracton Clinic, South East Thames | 63 | 74.6% | 33.6 | 63 | 100% | Home Office Offender Index | n/a | Case file reviews |
| Davison et al., 1999 | United Kingdom | Special Hospitals' Case Register | 159 | 57.4% | 34 | 50 | Unavailable | Home Office Offender Index | n/a | n/a |
| Friendship et al., 1999 | United Kingdom | The Denis Hill Unit at the Bethlem Royal Hospital, London | 234 | Unavailable | Unavailable | 159 | Unavailable | Home Office Offender Index | n/a | n/a |
| Falla et al., 2000 | United Kingdom | Trevor Gibbens Unit at Maidstone Hospital, Kent | 85 | Unavailable | Unavailable | 85 | Unavailable | Home Office Offender Index | Case file reviews | n/a |
| Halstead et al., 2001 | United Kingdom | The Eric Shepherd Unit (ESU) | 35 | 83% | 29.3 | 35 | 46% | Case file review | Case file review | n/a |
| Edwards et al., 2002 | United Kingdom | Three Bridges Unit, North West Thames | 225 | 85.4% | Unavailable | 104 | 79% | Home Office Offender Index | n/a | n/a |
| Dolan & Khawaja, 2004 | United Kingdom | Unnamed MSU, North West England | 70 | 100% | 44 | 70 | 73% | Home Office Offender Index | Case file review | n/a |
| Nicholls, 2004 | Canada | British Columbia Forensic Psychiatric Hospital (FPH) | 78 | 50% | 34.6 | 78 | 66.7% | British Columbia Correctional Movement sheets | n/a | British Columbia Division of Vital Statistics |
| Dowsett, 2005 | United Kingdom | Specialist Community | 47 | 94% | 37.7 | 47 | Unavailable | Case file review | n/a | n/a |
|  |  | Forensic Team,<br>Lambeth<br>Hospital |  |  |  |  |  |  |  |  |
| Seifert & Moller-Mussavi, 2005 | Germany | Essen<br>Prospective<br>Multicentre<br>Study | 333 | Unavailable | Unavailable | 333 | 50% | Federal<br>Central<br>Registry | n/a | n/a |
| Philipse et al.,<br>2006 | Netherlands | Pooled data<br>from 7 forensic<br>psychiatric<br>hospitals | 132 | 92.4% | 34.4 | 132 | 29.6% | Ministry of<br>Justice data | n/a | n/a |
| Simpson et al.,<br>2006 | New<br>Zealand | Auckland<br>Regional<br>Forensic<br>Psychiatry<br>Service | 105 | 87.6% | 37.5 | 48 | 86.7% | Case file<br>review | Case file<br>review | n/a |
| Coid et al., 2007 | United<br>Kingdom | Pooled data<br>collected from 7<br>of 14 medium<br>security forensic<br>services in<br>Thames | 1613 | 86.8% | 31.6 | 1344 | 62% | Home Office<br>Offender<br>Index | n/a | n/a |
| Akande et al.,<br>2007 | United<br>Kingdom | Unnamed low<br>secure unit,<br>Lewisham | 33 | 76% | 35.8 | 33 | 90% | Case file<br>review | Case file<br>review | n/a |
| Yoshikawa et al.,<br>2007 | Japan | Japanese<br>Ministry of<br>Justice registry | 489 | 84.3% | Unavailable | 489 | 67.3% | Japanese<br>Ministry of<br>Justice data | n/a | n/a |
| Blattner & Dolan,<br>2009 | United<br>Kingdom | Edenfield Centre | 72 | 87.5% | 36.4 | 39 | 67% | Home Office<br>Offender<br>Index | Case file<br>review | n/a |
| Fioritti et al., 2011 | Italy | Oidentary Hospital | 118 | Unavailable | Unavailable | 47 | Unavailable | Case file review | n/a | n/a |
| Nilsson et al., 2011 | Sweden | Unnamed forensic psychiatric hospital | 47 | Unavailable | Unavailable | 47 | Unavailable | National Council for Crime Prevention Register | n/a | n/a |
| Miraglia & Hall, 2011 | United States | Unnamed secure psychiatric facility, New York | 386 | 71.8% | 41.5 | 386 | Unavailable | New York State Division of Criminal Justice Services Criminal History database | n/a | n/a |
| Tabita et al., 2012 | Sweden | Unnamed medium secure forensic hospital, Örebro County | 88 | 90.9% | 40 | 88 | 31.8% | National Council for Crime Prevention Register | n/a | National Cause-of-Death Register |
| Marshall et al., 2014 | United States | Unnamed forensic hospital, Maryland | 356 | 78.1% | 39.6 | 356 | 71.3% | Maryland Office of Forensic Services case files | Maryland Office of Forensic Services case files | n/a |
| Salem et al., 2015 | Canada | Pooled data from 17 forensic hospitals in Québec | 837 | 82.4% | 35 | 837 | 64.5% | National police database | Case file review | n/a |
| Fazel et al., 2016 | Sweden | Swedish Patient Register | 6505 | 89.2% | 36.6 | 6505 | 33.6% | Swedish Crime Register | Swedish Patient Register | National Cause-of-Death Register |
| Simpson et al., 2018 | Canada | Canadian Provincial Review Boards discharged patient data | 60 | 85% | 44.5 | 60 | 81.7% | Case file review | Case file review | n/a |
| Penney et al., 2018 | Canada | Data collected from 2 unnamed forensic psychiatric hospitals | 87 | 83.9% | 36.4 | 87 | 81.6% | n/a | Case file review | n/a |
| Jewell et al., 2018 | United Kingdom | South London and Maudsley NHS Foundation Trust forensic inpatient services | 101 | 82.2% | 40 | 101 | 89.1% | n/a | Case file review | n/a |
| deVogel et al., 2019 | The Netherlands | Data collected from 4 forensic hospitals | 78 | 0% | 38.6 | 78 | Unavailable | Ministry of Justice and Security judicial documentation register | Case file review | Case file review |
| Hogan et al., 2019 | Canada | Unnamed maximum-security hospital | 82 | 93.9% | 37.7 | 82 | 75.6% | Case file review | n/a | n/a |
| Bengtson et al., 2019 | Denmark | Data collected from 2 unnamed forensic hospitals in Copenhagen and Aarhus | 392 | 95.4% | 32.7 | 392 | 57.4% | Danish Crime Register | n/a | n/a |
| Nagata et al., 2019 | Japan | Data collected from 28 of 30 Forensic Medical Treatment and Supervision Act facilities | 526 | 79.5% | Unavailable | 526 | 82.3% | Case file review | Case file review | n/a |
| Ojansuu et al., 2019 | Finland | Finnish National Institute for Health and Welfare Database | 950 | 86.6% | 43 | 950 | 57.1% | n/a | n/a | National Cause-of-Death Register |
| Takeda et al., 2019 | Japan | Data collected from 29 of 33 forensic psychiatric wards throughout Japan. | 966 | 76.5% | 47.3 | 966 | 81.3% | n/a | n/a | Case file review |
| Probst et al., 2020 | Germany | Data collected from 2 unnamed forensic psychiatric clinics in Bavaria | 249 | Unavailable | Unavailable | 249 | Unavailable | Case file review | n/a | n/a |
| Rossetto et al., 2021 | Italy | Residences for the Execution of the Security Measure of Castiglione delle Stiviere | 90 | 0% | 38.2 | 90 | 47.8% | n/a | Case file review | n/a |
| Dean et al., 2021 | Australia | NSW Forensic hospital* | 477 | 86.2% | 33.8 | 270 | 83.9% | NSW Bureau of Crime Statistics and Research (BOCSAR) Re-offending database | n/a | n/a |
| Lyons et al., 2022 | Australia | NSW Forensic hospital* | 477 | 86.2% | 33.8 | 245 | 83.9% | n/a | Case file review | n/a |
| Fovet et al., 2022 | France | French National Hospital Database | 3020 | 88.8% | Unavailable | 1053 | 61.8% | n/a | French national hospital databas | n/a |
| Haroon et al., 2023 | United States | North Carolina Forensic Treatment Program Database | 61 | 82% | 36.3 | 61 | 68.9% | North Carolina Public Offender database | Hospital patient directory |  |
| Ojansuu et al., 2023 | Finland | Finnish National Institute for | 501 | 86.6% | 47 | 501 | 89.6% | Institute of Criminology | n/a | n/a |
|  |  | Health and Welfare registry |  |  |  |  |  | and Legal Policy database |  |  |
| Argo et al., 2023 | Israel | Israeli Ministry of Health's National Psychiatric Hospitalization Registry | 136 | 92.6% | Unavailable | 136 | 79.4% | n/a | National Psychiatric Hospitalization Registry | n/a |
| Sivak et al., 2023 | Sweden | Swedish National Forensic Psychiatric Register (SNFPR) | 657 | 79.4% | 42.3 | 640 | 70.6% | Swedish National Council for Crime Prevention's crime registry | n/a | n/a |
| Thomson & Rees, 2023 | United Kingdom | State Hospital Survey (SHS) for patients detained in high secure state hospitals in Scotland | 241 | 56.4% | 37.4 | 151 | 62.7% | Scotland Police data | n/a | n/a |
| Westhead et al., 2023 | United Kingdom | Arnold Lodge, England | 909 | 84.4% | 31.7 | 521 | 51.9% | Police National Computer (PNC) by the Ministry of Justice (MoJ) | Office for National Statistics (ONS) and Hospital Episode Statistics (HES) data | Office for National Statistics (ONS) and Hospital Episode Statistics |

|  |  |  |  |  |  |  |  |  |  | (HES)<br>data |
| --- | --- | --- | --- | --- | --- | --- | --- | --- | --- | --- |
| Hill et al., 2024 | United Kingdom | Patients discharged from 35 NHS and independent sector medium secure units | 616 | 88.2% | Unavailable | 164 | Unavailable | Home Office Offender Index | n/a | n/a |

\* = overlapping dataset that reported on different outcomes, SSD = Schizophrenia Spectrum Disorders.

Of the studies meeting the inclusion criteria, the majority (78%) were rated as high quality (*n =* 38). A smaller proportion (22%) were classified as moderate (*n* = 11) and none were considered low quality. The total scores for each study are shown in the corresponding forest plots (See Figures 2 – 7).

### Any Reoffending

Thirteen studies (*n* = 1,883) reported the rates of any reoffending among patients discharged from secure care ^2,17,19,40–49^. A random-effects model showed that crude reoffending rates ranged from 275 to 16,667 per 100,000 person-years, with a pooled estimate of 3,889, 95% CI [2,055, 7,359], *I^2^* = 97%, 95% CI [96% - 97.8%] (see Figure 2). In terms of heterogeneity, the 95% prediction interval ranged from 290 to 52,136, indicating substantial variability in the effect sizes between studies.

**Figure 2.**
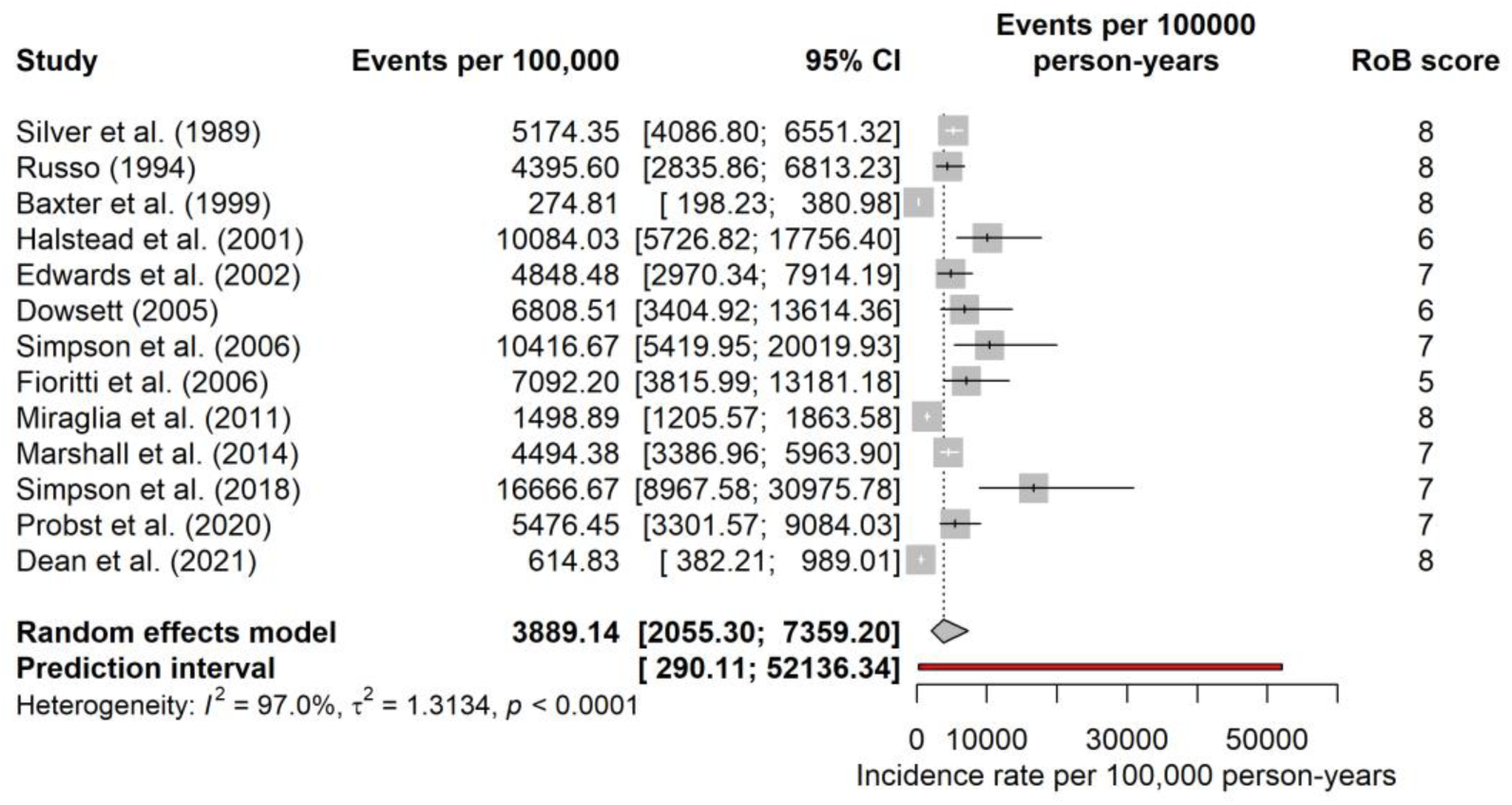
Forest plot of any reoffending rates among forensic patients with community access expressed as per 100,000 person-years. Pooled estimates calculated using a Random-effect model. Red line denotes the 95% prediction interval.

### Violent Reoffending

Twenty-eight studies (*n* = 12,764) provided data on violent reoffending rates among patients discharged from secure care ^2,4,17,19,20,22,24,41,42,44,45,47,50–65^. A random-effects model showed that crude violent reoffending rates range from 145 to 16,667 per 100,000 person-years, with a pooled estimate of 1,851, 95% CI [1,229, 2,789], *I^2^* = 95.3%, 95% CI [94.1%-96.2%] (see Figure 3). In terms of heterogeneity, the 95% prediction interval ranged from 201 to 17,068 indicating substantial variability in the effect sizes between studies.

**Figure 3.**
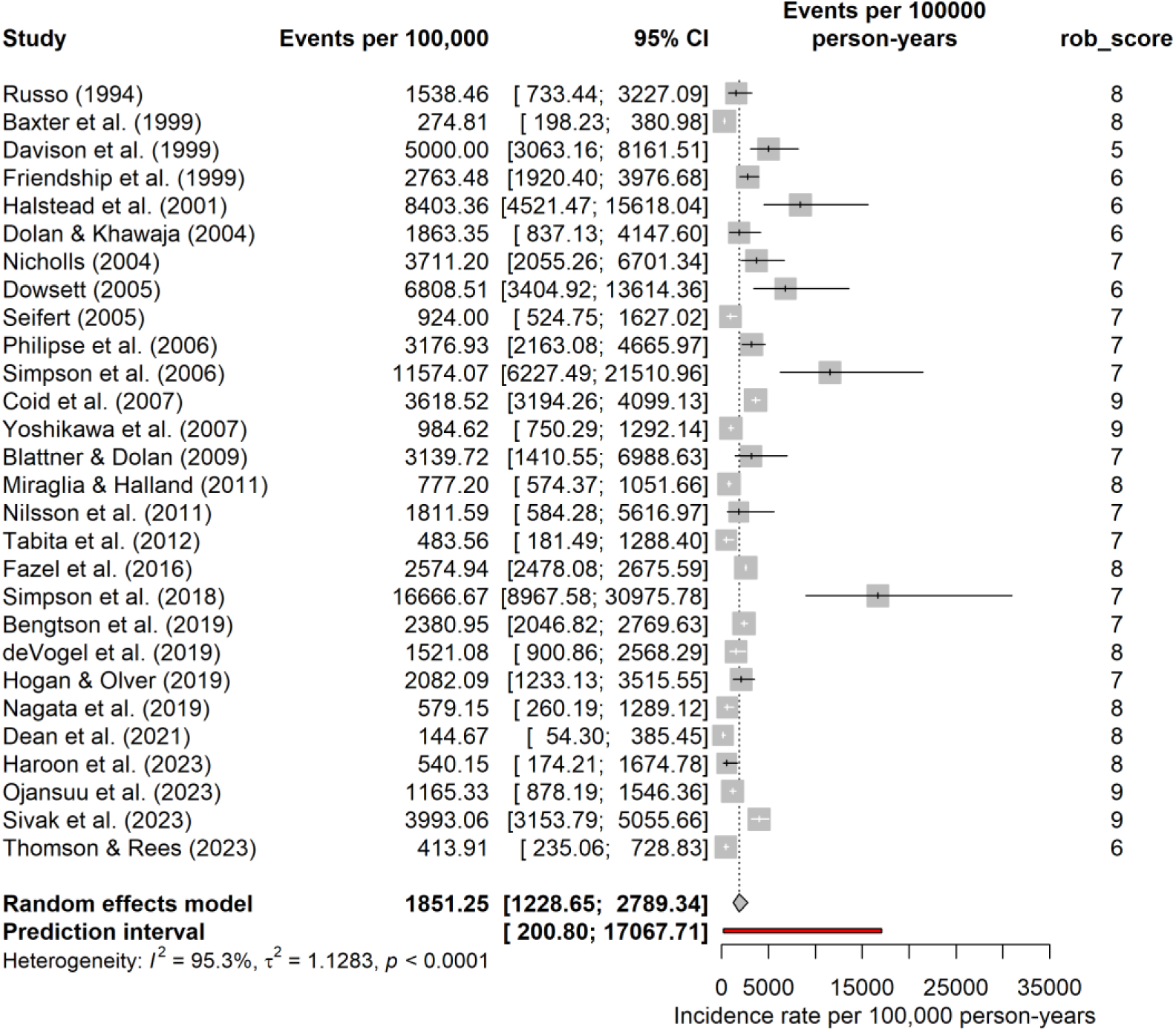
Forest plot of violent reoffending rates among forensic patients with community access expressed as per 100,000 person-years. Pooled estimates calculated using a Random-effect model. Red line denotes the 95% prediction interval.

### Reconvictions

Twenty-nine studies (*n* = 13,327) reported on all reconviction rates among patients discharged from secure care ^1,2,16,18–20,22,24,42,43,45,50–52,54,55,57–69^. A random-effects model showed that crude reconviction rates range from 0 to 7,733 per 100,000 person-years, with a pooled estimate of 3,291, 95% CI [2,591, 4,179], *I^2^* = 96.2%, 95% CI [95,3% - 96.9%] (see Figure 4). In terms of heterogeneity, the 95% prediction interval ranged from 950 to 11,394, indicating substantial variability in the effect sizes between studies.

**Figure 4.**
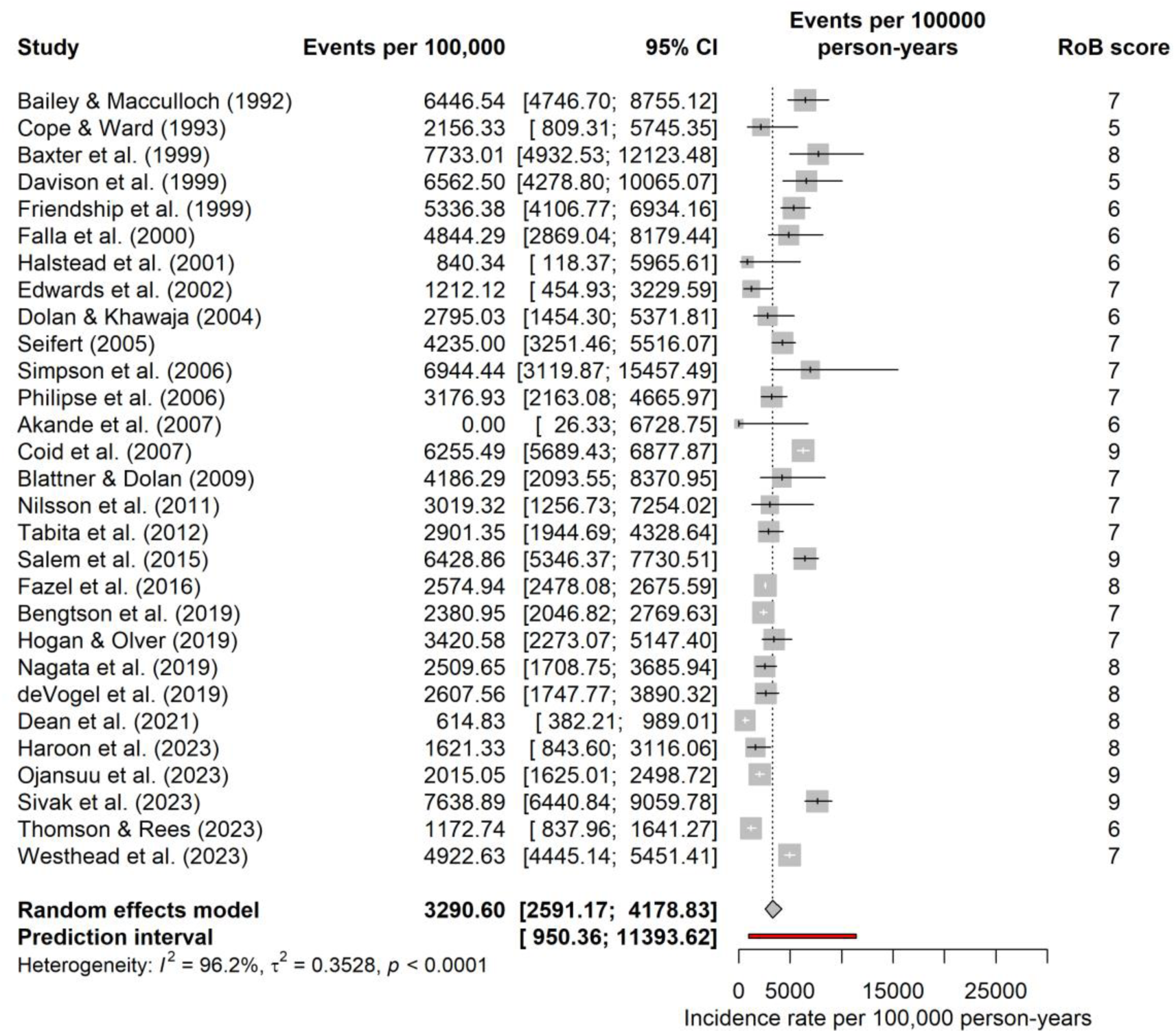
Forest plot of reconviction rates among forensic patients with community access expressed as per 100,000 person-years. Pooled estimates calculated using a Random-effect model. Red line denotes the 95% prediction interval.

### Readmissions

Twenty-four studies (*n* = 11,228) reported on readmission rates among patients discharged from secure care ^1,4,18,19,21,23,42,45,47,49,52,57,60–62,67–75^. A random-effects model showed that crude readmission rates range from 978 to 27,586 per 100,000 person-years, with a pooled estimate of 7,945, 95% CI [5,507, 11,463], *I^2^*= 98.9%, 95% CI [98.8% - 99.1%] (see Figure 5). In terms of heterogeneity, the 95% prediction interval ranged from 1,225 to 51,548, suggesting substantial variability in the effect sizes between studies.

**Figure 5.**
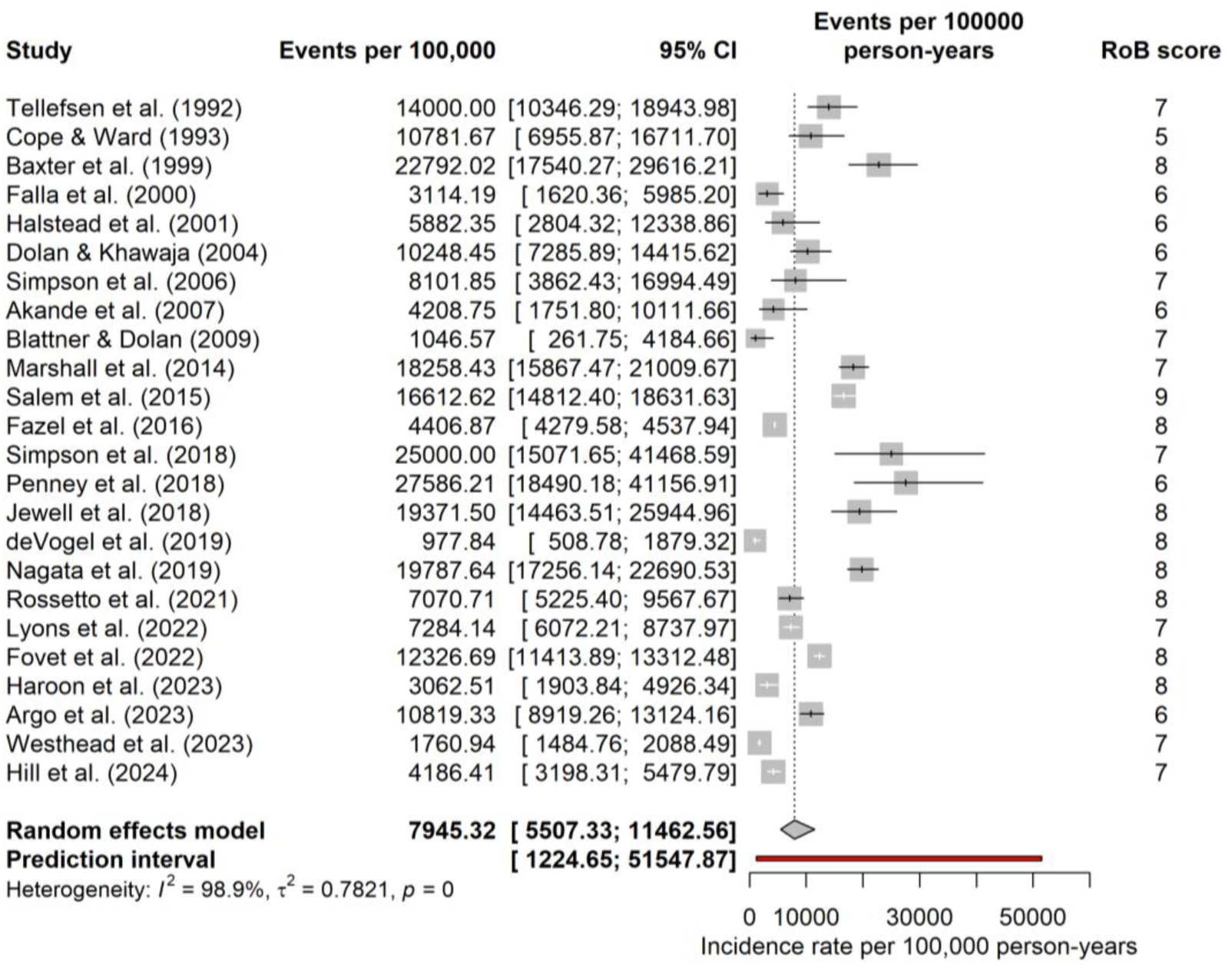
Forest plot of readmission rates among forensic patients with community access expressed as per 100,000 person-years. Pooled estimates calculated using a Random-effect model. Red line denotes the 95% prediction interval.

### All-cause Mortality

Ten studies (*n* = 9,564) reported on all-cause mortality rates among patients discharged from secure care ^1,4,20,41,53,60,65,70,76,77^. A random-effects model showed that crude all-cause mortality rates range from 735 to 3,173 per 100,000 person-years, with a pooled estimate of 1,789, 95% CI [1,341, 2,388], *I^2^* = 89.4%, 95% CI [82.7% - 93.6%] (see Figure 6). In terms of heterogeneity, the 95% prediction interval ranged from 673 to 4,756, indicating substantial variability in the effect sizes between studies. Meta-regression was not conducted due to missing data on follow-up duration in one study, resulting in fewer than 10 studies that had complete information.

**Figure 6.**
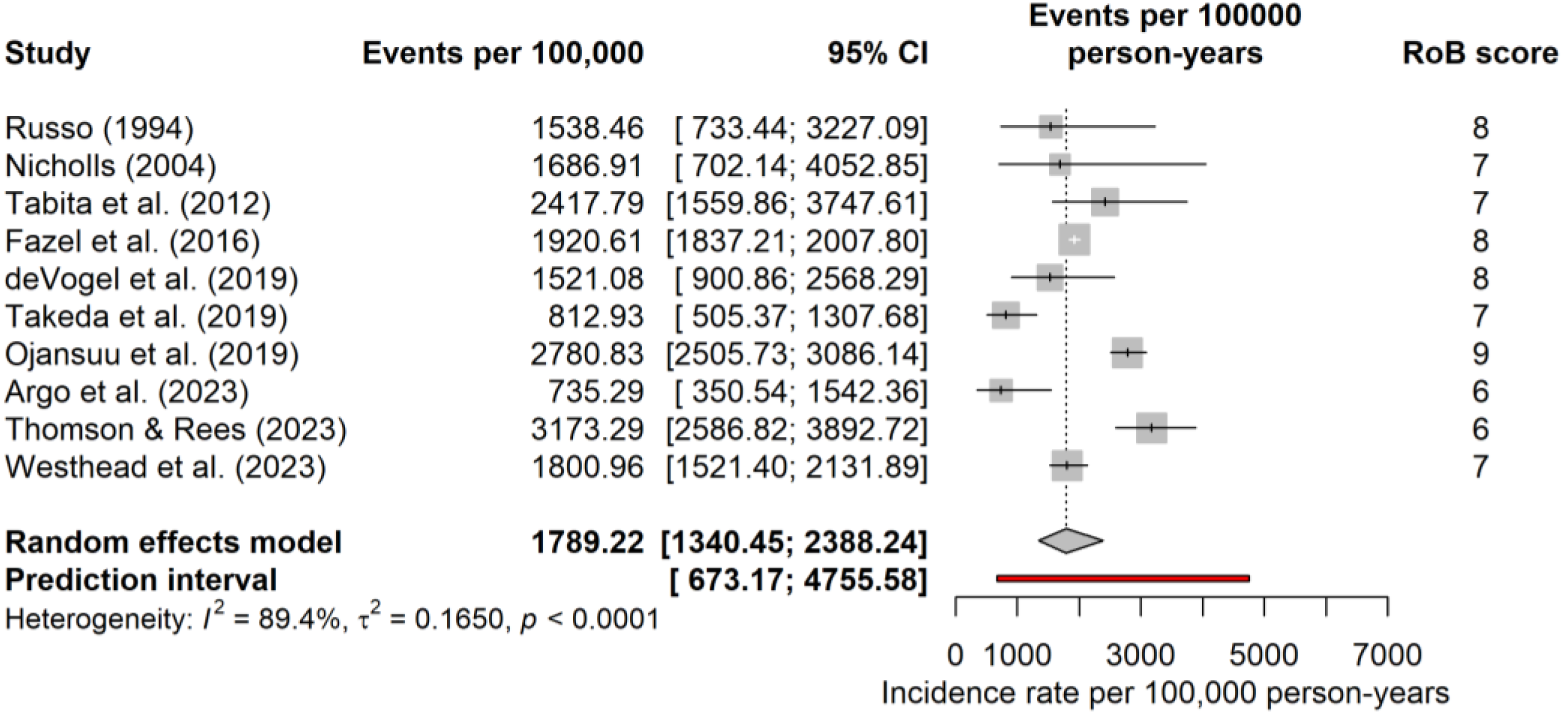
Forest plot of all-cause mortality rates among forensic patients with community access expressed as per 100,000 person-years. Pooled estimates calculated using a Random-effect model. Red line denotes the 95% prediction interval.

### Completed Suicide

Eight studies (*n* = 9,335) reported on suicide rates among patients discharged from secure care ^1,4,20,41,60,70,76,77^. A random-effects model showed that crude suicide rates range from 210 to 725 per 100,000 person-years, with a pooled estimate of 407, 95% CI [319, 519], *I^2^* = 41.5%, 95% CI [0.0% - 74.2%] (see Figure 7). In terms of heterogeneity, the 95% prediction interval ranged from 225 to 735, indicating minimal variability in the effect sizes across individual studies.

**Figure 7.**
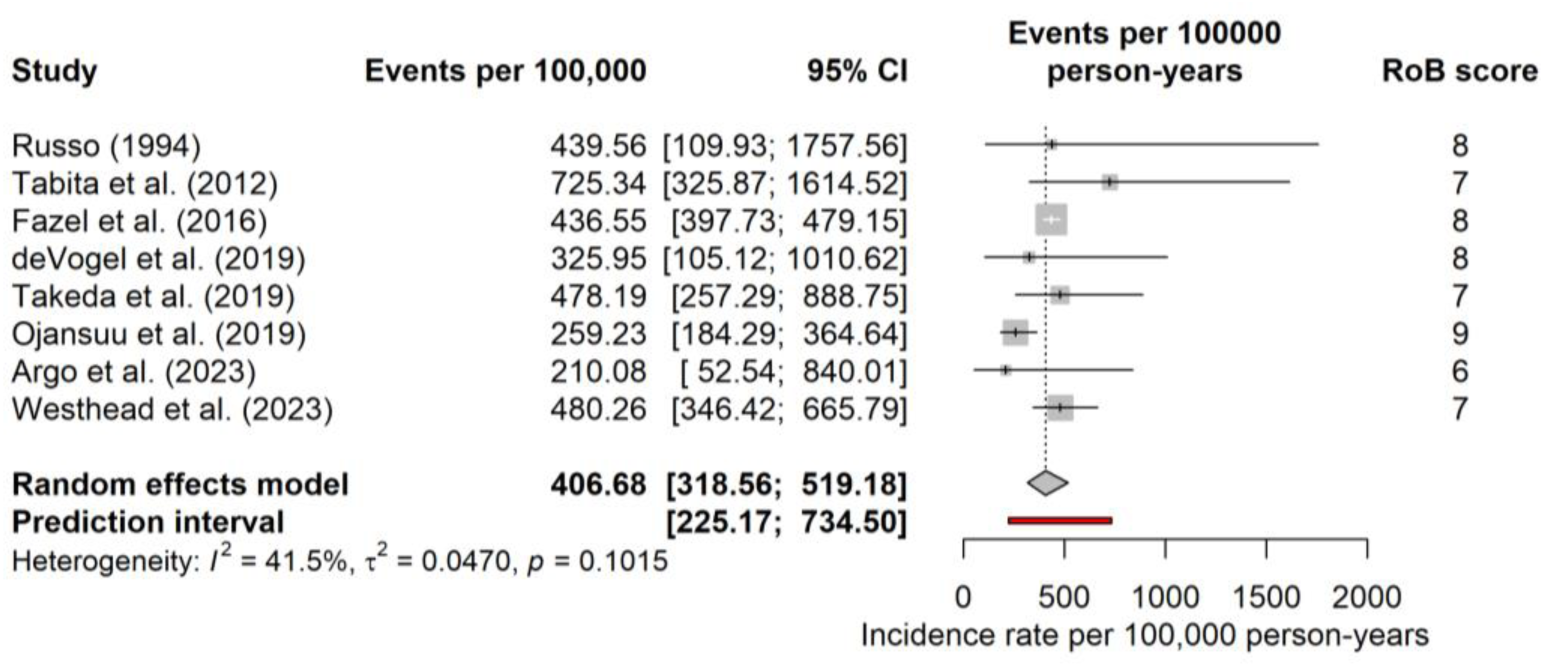
Forest plot of suicide rates among forensic patients with community access expressed as per 100,000 person-years. Pooled estimates calculated using a Random-effect model. Red line denotes the 95% prediction interval.

### Moderator Analyses

Meta-regression analyses were conducted to explore the effects of mean follow-up duration and proportion of patients with schizophrenia accounted for between study heterogeneity across outcomes.

For any reoffending, there was a negative but non-significant association with mean follow-up duration, *B* = -0.14, *SE* = 0.08, *z* = -1.85, *p* = .07, 95% CI [-0.28, 0.01]. This finding suggests that studies with longer follow-up periods tended to report lower any reoffending rates, explaining approximately 17% of the between-study variance (*R*^2^ = 16.53%).

For violent reoffending, there was a significant negative correlation with mean follow-up duration, *B* = -0.09, *SE* = 0.04, *z* = -2.1, *p* = .04, 95% CI [-0.1, 0.01]. This indicates that studies with longer follow-up periods were significantly more likely to report lower rates of violent reoffending, accounting for approximately 12% of between-study heterogeneity (*R*^2^ = 12.3%). In contrast, the proportion of patients with schizophrenia was not associated with violent reoffending, *B* = -0.01, *SE* = 0.01, *z* = -0.8, *p* = .426, 95% CI [-0.03, - 0.01], *R^2^* = 0%.

For reconvictions, there was a significant negative association with mean follow-up duration, *B* = -0.05, *SE* = 0.02, *z* = -2.43, *p* = .02, 95% CI [-0.1, -0.01], *R*^2^ = 22.40%, but not with the proportion of patients with schizophrenia, *B* = -0.003, *SE* = 0.01, *z* = -0.43, *p* = .67, 95% CI [-0.02, 0.01], *R*^2^ = 0%. The moderator analysis suggest study duration was inversely associated with reconvictions, with longer follow-up periods linked to lower reported rates.

For readmissions, both moderates showed significant effects when run as independent meta-regression models. Mean follow-up duration was negatively associated with readmission rates, *B* = -0.15, *SE* = 0.04, *z* = -4.17, *p* < .001, 95% CI [-0.21, -0.08], *R*^2^ = 47.42%. In contrast, the proportion of patients with schizophrenia was positively associated with readmissions, *B* = 0.02, *SE* = 0.01, *z* = 3.26, *p* < .001, 95% CI [-0.01, -0.04], *R*^2^ = 56.93%. In the combined model with both moderators, however, only mean follow-up duration retained statistical significance, *B* = -0.09, *SE* = 0.03, *z* = -2.86, *p* = .004, 95% CI [-0.14, -0.03], while proportion of schizophrenia did not, *B* = 0.008, *SE* = 0.007, *z* = 1.15, *p* = .25, 95% CI [-0.01, 0.02]. Similar to previous outcomes, these findings indicate an inverse association between follow-up time and readmission incidences.

## Discussion

Our meta-analysis synthesised evidence on six key health and justice outcomes among forensic patients discharged from secure settings into the community. Across 49 studies published up to May 2025, pooled incidence rates per 100,000 person-years were calculated for all reoffending, violent reoffending, reconvictions, psychiatric readmissions, all-cause mortality, and suicide. These studies included a total of 18,871 patients across 14 countries, contributing to approximately 197,114 total person-years of observation. Overall, the highest observed pooled incidence rate was psychiatric readmission, followed by reoffending outcomes. While suicide and all-cause mortality occurred less frequently, they remain elevated among forensic patients in the community. There were substantial between-study heterogeneity across outcomes, suggesting meaningful variation in incidence risk depending on factors such as research methodologies and patient characteristics.

### Reoffending Outcomes

The pooled incidence rates for discharged forensic patients were 3,889 for any reoffending, 1,851 for violent reoffending, and 3,291 for reconviction per 100,000 person-years. Overall, these findings suggest that patients discharged from secure care are, broadly speaking, reoffending less than other justice involved populations. Equivalent justice outcomes for general population comparison were obtained from the Bureau of Statistics and Research^78^ (BOCSAR) crime registry in New South Wales, Australia. The BOCSAR data is one of the only sources of comparable publicly available data, drawing on a large cohort of 93,550 previously convicted individuals over 10 years. The pooled violent reoffending rate for forensic patients discharged to the community was, for instance, approximately half of that reported in the most recent BOCSAR report for people previously charged with a violent index offence (3,900 per 100,000 person-years) ^78^. Similarly, the pooled conviction rate of forensic patients was 3,291 per 100,000 person-years, substantially lower than for people previously convicted of any offences, at 5,355 per 100,000 person-years^78^. While these findings are encouraging, it should be noted, however, that secure care patients differ in many key respects from other offender groups. Forensic psychiatric patients, for example, are more likely to be charged with serious violent offences (e.g. high proportions charged with homicide offences), experience lengthier periods of detention, and closer monitoring upon discharge^79^. Despite these differences, the consistent lower violent reoffending rates support the notion that treatment and rehabilitation focused on the primary drivers of violence risk for those with mental illness, may ultimately be successful in reducing such risk.

Additionally, notable differences in outcomes were identified when compared to the previous Fazel et al.^16^ review of discharged forensic patients. In the Fazel et al. ^16^ study, the pooled violent reoffending rate was 3,902 per 100,000 person-years. In the current review, restricted to studies of forensic patients discharged to the community covering the same period (i.e., 1982-2013) and extending to 2025, the violent reoffending rate was much lower at 1,851, per 100,000 person-years. One explanation is that advances in forensic psychiatric care and stricter post-discharge monitoring have together contributed to a lower reoffending rate in recent decades. To examine this, we performed a post-hoc sensitivity analysis excluding studies published prior to the Fazel et al.^16^ review search period (i.e., only studies published after 2013, *n* = 11). The analysis produced a pooled *violent reoffending* estimate of 1,416 per 100,000 person-years, approximately 65% lower than the equivalent rate in the Fazel et al ^16^ study (see Appendix A). Similarly, when restricted to studies after 2013, the pooled *reconviction rates* were about 40% lower, at 2,612 per 100,000 person-years, compared to Fazel et al.’s ^16^ reported rate of 4,484 per 100,000 person-years (see Appendix A). This decline supports the notion that contemporary forensic services are more effective at addressing the recidivism risk factors amongst those with severe mental illnesses, leading to improved justice outcomes. However, without a direct comparison to a non-forensic offender group over the same time period, the possibility of cohort trends in repeat offending cannot be ruled out.

Our meta-regression model showed that that studies with longer follow-up tended to report lower reoffending rates. This is consistent with the broader recidivism literature demonstrating that reoffending is more likely to occur shortly after release ^80^. Thus, as with other offender groups, forensic patients may be at greater risk of reoffending soon after discharge, with this risk diminishing gradually overtime, a phenomenon that has important implications for clinical care. It is worth noting, however, that this effect can only be systematically assessed if studies report follow-up data in a more standardised manner (e.g., at 12-months, 1-year, 3-year, 5-year, and 10-year). Adopting a standardised approach in future research will enable for more robust sensitivity analyses in future meta-analytic investigations.

### Psychiatric Readmission

One of the key health outcomes examined was the psychiatric readmission rates among forensic patients discharged from secure care to the community. The pooled estimated rate of psychiatric readmission was 7,945 per 100,000 person-years, which is similar to that reported in the previous meta-analysis conducted^16^ over a decade earlier at 7,208 per 100,000 person-years. However, when restricting analyses to studies published within the past 10 years since the Fazel et al. ^16^ review, a slightly higher rate of readmission for the more recent period was identified, approximately 8,314 per 100,000 person-years (see Appendix A in Supplementary material). The disparity between the study periods suggests that, despite potential improvements in forensic psychiatric care, patients are returning to hospital more frequently than previously. This may reflect appropriate clinical intervention resulting from careful monitoring of discharged forensic patients rather than an adverse outcome to be prevented at all costs. Serious mental illnesses like schizophrenia typically have a relapsing course ^81^, despite optimal treatment and support in severe cases, and the readmission rates likely reflect this reality, at least in part.

The upward trend in rehospitalisation rates may also be due to recent studies relying on more comprehensive and reliable data sources (e.g., nation-wide electronic health registries) than those conducted in previous decades. Nevertheless, the elevated readmission rates in more recent studies warrants closer examination. Future investigations should consider recording greater detail about the reasons for readmission to clarify the post-discharge challenges forensic patients face to guide more targeted intervention. Importantly, on meta-regression analysis, SSD prevalence largely accounted for the variability in readmissions between studies, with studies including a greater proportion of forensic patients with SSD reporting higher rehospitalisation rates. This pattern aligns with findings from general psychiatric population studies in which patients with SSD diagnoses are readmitted more frequently than those diagnosed with other mental illness diagnoses ^82^.

### All-cause Mortality and Suicide

The pooled all-cause mortality rate for forensic patients discharged to the community was 1,789 per 100,000 person-years. This rate was twice that reported in a meta-analysis of former offenders released from prisons based in the United States, UK, and Australia, at 850 per 100,000 person-years ^83^. Although higher than that of former prisoners, the all-cause mortality rate for forensic patients appears only slightly above estimates reported for adults with schizophrenia in the United States, around 1,540 per 100,000 person-years. Together, these comparisons indicate that forensic patients are may be dying at a greater rate than former offenders but show a similar mortality pattern to general psychiatric patients.

The high all-cause mortality rate among forensic patients is likely due to a confluence of the impact of lifestyle, treatment effects, and systematic factors that undermine their ability to maintain a healthy life. People with severe mental illness, like schizophrenia, are more susceptible to unhealthy behaviours, such as smoking ^84^ and substance misuse ^85^, both of which dramatically increase the likelihood of premature death. Obesity is another major concern for patients with severe mental illness, including as a result of side effects arising from treatment with antipsychotic medication, because of its impact on risk of cardiovascular disease, diabetes mellitus, and cancers ^86–88^. Additionally, negative symptoms of schizophrenia, along with sedation arising from antipsychotic treatment, may lead to reduced motivation for physical activity ^89,90^. Living in secure institutional settings which often offer limited access to outdoor spaces to maintain an active lifestyle can also exacerbate the problem ^91^. Finally, there are also post-discharge barriers that are associated with physical health inequalities, including difficulty accessing preventative healthcare (e.g., being referred for cancer screening) and stigma from the broader community ^92^.

The pooled rate of deaths by suicide for forensic patients discharged from secure care to the community was 407 per 100,000 person-years. This rate is similar to that reported in other high-risk psychiatric populations and to the previous review of forensic patients^16^. For instance, Chung et al.’s ^93^ meta-analysis of 100 studies of discharged general psychiatric patients reported a suicide incidence rate of 484 per 100,000 person-years. The pooled estimate for the suicide rate for forensic patients also aligns with that of community patients with SSD, at 441 per 100,000 person-years ^94^. These similarities suggest that, despite lengthier and more resource-intensive treatment, forensic patients discharged from secure care remain equally vulnerable to suicide. This likely reflects the reality that forensic patients, in addition to schizophrenia, typically present with other co-morbidities that may complicate their suicide risk, including substance use problems, personality disorders, and cognitive impairments ^2^. Moreover, forensic mental health services have historically prioritised managing the patients’ risk to others and protecting the broader community, occasionally through contentious restrictive practices, including restraints and seclusions^95^. This emphasis on external risks may have contributed to the neglect of an equally important concern: patients’ risk of self-harm and suicide. Hence, these results reaffirm the need for service providers to recognise the heightened suicide risk in this patient group and invest more into community-based monitoring and preventative measures.

## Strengths and Limitations

The current systematic review and meta-analysis has several notable strengths. One major strength of the meta-analysis is that it only included forensic patients that were discharged to the community, and thus had spent time at risk of committing a crime or be readmitted. Previous reviews have included studies with a proportion of patients transferred to other secure facilities, inpatient psychiatric units or to prisons. Restricting inclusion to only those cohorts with confirmed time at risk produces incidence rates more representative of the forensic and clinical challenges discharged patients encounter in the community. Additionally, we chose not to impose a restriction on language on the database searches, which allowed us to include two additional estimates from non-English speaking countries. It is imperative to include studies published in other languages in these global estimates as a majority of the data tend to come from Western, English-speaking countries. Finally, the review covered an extended search period, from start of digitised databases through to 2025. This broader timeframe allowed us to examine differences between time and generate pooled estimates for more specific outcome types (e.g., reconvictions) that have yet been reported meta-analytically.

There are, however, important limitations in the existing studies that must be taken into consideration. First, nearly half of the studies included in our analysis were likely underpowered, with 23 out of the 49 studies including a sample with fewer than 100 patients. There are statistical corrections that can be applied to reduce the influence of small-study effects ^96^. However, given incidence studies have no true zero, we could not adequately assess the extent to which small-study effects influenced our findings.

The high levels of heterogeneity and the wide prediction interval range identified across studies, with the exception of suicide rates, indicated substantial between-study variation. While follow-up duration accounted for some of this heterogeneity, the number of eligible studies were insufficient to further examine other potential moderators. The variability in findings likely reflect a complex interplay of factors including differences in study design, national health polices, and jurisdictional definitions of forensic patients. Future secondary analyses, where a greater number of studies can be included, might benefit from region-specific synthesis of incidence rates to provide more relevant and accurate estimates.

Moreover, moderator analyses were limited for two reasons. First, the number of eligible studies per outcome was often smaller than anticipated. As recommended in the Cochrane Handbook for Systematic Reviews of Interventions ^39^, a ratio of at least 10 observations per moderator is preferred to avoid overfitting in meta-regression models. Hence, only one or two moderators were used in most outcomes. The second problem was that studies often report demographic information inconsistently across subsamples, making it difficult to extract all the relevant characteristics required for meta-regressions. These factors limited our ability to explore potential moderators and potentially contributed to the unexplained heterogeneity across studies.

Finally, most studies failed to report the total person-time data needed to calculate incidence rates. Given less than 10 percent of the papers (*n* = 6) reported total time at risk, crude estimates were calculated using the mean follow-up duration. However, as Clark et al. ^34^ recently demonstrated, crude estimates may not adequately account for unpredictable patterns in follow-up data. Clark et al. ^34^ noted that out of the five approaches tested, the pseudo-individual participant data (IPD) approach calculated using the Kaplan-Meier curves yielded the most robust results. Unfortunately, it was not possible to adopt this approach as only 14 studies reported the required curves. Given the importance of research synthesis in shaping policies and practice, future studies should aim to provide enough sufficient detail to support secondary analyses, including definitions of outcomes defined, sources of information (e.g., official vs unofficial records), and total person-year data. Doing so will help strength the validity of future meta-analytic work in this field.

## Implications for Practice and Research

There are several important implications to draw from these findings. First, although many forensic patients have previously committed serious violent offences, our analyses suggests that their reoffending rates once returned to the community are lower than for non-forensic individuals with previous criminal history, including those committing violent offences. Another implication is that, despite their lengthier and more resource intensive treatment, forensic patients appear to be just as likely to die prematurely as their general psychiatric counterparts. Their premature death rate also far exceeds both people in the general population and people released from prison, pointing to a clear disparity between forensic patients and others in the community. Additionally, our results show that, despite intensive care regimes in many secure settings, patients’ suicide risk remains high compared to non-forensic populations and is comparable to other high-risk psychiatric patients. This suggests it is vital for forensic services, particularly in post-discharge care, to consider risks of harm not only to the broader community, but also to the patients themselves.

## Supporting information

Supplementary Analyses

## Author contributions

J.M. contributed to the conceptualisation, question formulation, systematic screening of papers, data extraction, data analysis, and writing the original draft manuscript. C.M. contributed to the conceptualisation, question formulation, systematic screening of papers, data extraction, data analysis and editing the manuscript. R.K. contributed to the conceptualisation, question formulation, and editing the manuscript. K.D. contributed to the conceptualisation, question formulation, and editing the manuscript.

## Funding

The first author, JM, is a PhD candidate supported by an Australian Government Research Training Program Scholarship. RK is partially supported by ARC funding (DP250102628, DP220100585 and LP230201076). KD is supported by the NHMRC Investigator Grant (APP1175408) and Justice Health NSW.

## Transparency declaration

J.M. declares that this manuscript is an honest, accurate, and transparent account of the study being reported. No important aspects of the study have been omitted, and any discrepancies from the study as planned have been explained.

## Declaration of interests

The authors declared no conflict of interests

## Data Availability

The data used to in this study are available upon requestion.

## Notes

### Competing Interest Statement

The authors have declared no competing interest.

### Author Declarations

This systematic review and meta-analysis was completed based on publicly available studies.

## References

1 Westhead J, Gibbon S, McCarthy L, Hatcher R, Clarke M. Long-term outcomes after discharge from medium secure care: still a cause for concern? J Forensic Psychiatry Psychol 2023; 34: 166–78.

2 Dean K, Singh S, Kemp R, Johnson A, Nielssen O. Characteristics and re-offending rates amongst individuals found not guilty by reason of mental illness (NGMI): A comparison of men and women in a 25-year Australian cohort. Int J Forensic Ment Health 2021; 20: 17–30.

3 Whiting D, Gulati G, Geddes JR, Fazel S. Association of schizophrenia spectrum disorders and violence perpetration in adults and adolescents from 15 countries: A systematic review and meta-analysis. JAMA Psychiatry 2022; 79: 120–32.

4 Fazel S, Wolf A, Fimiñska Z, Larsson H. Mortality, rehospitalisation and violent crime in forensic psychiatric patients discharged from hospital: Rates and risk factors. PLOS ONE 2016; 11: 1–14.

5 Waqar MU, Amin H, Ní Mhuircheartaigh E, Kennedy HG, Davoren M. Prevalence of Treatment Resistant Psychoses in a Complete National Forensic Mental Health Service: A Dundrum forensic redevelopment evaluation study (D-FOREST). Eur Psychiatry 2023; 66: 431.

6 Melnychuk RM, Verdun-Jones SN, Brink J. Geographic risk management: A Spatial study of mentally disordered offenders discharged from forensic psychiatric care. Int J Forensic Ment Health 2009; 8: 148–68.

7 Gu Y, Guo H, Zhou J, Wang X. Socio-demographic, clinical and offense-related characteristics of forensic psychiatric inpatients in Hunan, China: A cross-sectional survey. BMC Psychiatry 2023; 23: 48: 1–9.

8 Landgraf S, Blumenauer K, Osterheider M, Eisenbarth H. A clinical and demographic comparison between a forensic and a general sample of female patients with schizophrenia. Psychiatry Res 2013; 210: 1176–83.

9 Ekinci O, Ekinci A. Association between insight, cognitive insight, positive symptoms and violence in patients with schizophrenia. Nord J Psychiatry 2013; 67: 116–23.

10 Darrell-Berry H, Berry K, Bucci S. The relationship between paranoia and aggression in psychosis: A systematic review. Schizophr Res 2016; 172: 169–76.

11 Fazel S, Gulati G, Linsell L, Geddes JR, Grann M. Schizophrenia and violence: Systematic review and meta-analysis. PLOS Med 2009; 6: 1–15.

12 Yee N, Matheson S, Korobanova D, Large M, Nielssen O, Carr V, et al. A meta-analysis of the relationship between psychosis and any type of criminal offending, in both men and women. Schizophr Res 2020; 220: 16–24.

13 Völlm B, Bartlett P, McDonald R. Ethical issues of long-term forensic psychiatric care. Ethics Med Public Health 2016; 2: 36–44.

14 Rutherford M, Duggan S. Forensic mental health services: Facts and figures on current provision. Br J Forensic Pract 2008; 10: 4–10.

15 Australian Institute of Health and Welfare. Health Expenditure Australia 2022–23. Australian Institute of Health and Welfare, 2024.

16 Fazel S, Fimiñska Z, Cocks C, Coid J. Patient outcomes following discharge from secure psychiatric hospitals: Systematic review and meta-analysis. Br J Psychiatry 2016; 208: 17–25.

17 Miraglia R, Hall D. The effect of length of hospitalization on re-arrest among insanity plea acquittees. J Am Acad Psychiatry Law 2011; 39: 524–34.

18 Akande E, Beer MD, Ratnajothy K. Outcome study of patients exhibiting challenging behaviours four years after discharge from a low secure mental health unit. J Psychiatr Intensive Care 2007; 3: 21–6.

19 Halstead S, Cahill A, Fernando L, Isweran M. Discharges from a learning-disability medium secure unit: What happens to them? Br J Forensic Pract 2001; 3: 11–21.

20 Tabita B, de Santi, Miguel G., and Kjellin L. Criminal recidivism and mortality among patients discharged from a forensic medium secure hospital. Nord J Psychiatry 2012; 66: 283–9.

21 Penney SR, Marshall L, Simpson AIF. A prospective study of pathways to hospital readmission in Canadian forensic psychiatric patients. J Forensic Psychiatry Psychol 2018; 29: 368–86.

22 Hogan NR, Olver ME. Static and dynamic assessment of violence risk among discharged forensic patients. Crim Justice Behav 2019; 46: 923–38.

23 Fovet T, Baillet M, Horn M, Chan-Chee C, Cottencin O, Thomas P, et al. Psychiatric hospitalizations of people found not criminally responsible on account of mental disorder in France: A ten-year retrospective study (2011–2020). Front Psychiatry 2022; 13: 2–8.

24 Coid J, Hickey N, Kahtan N, Zhang T, Yang M. Patients discharged from medium secure forensic psychiatry services: Reconvictions and risk factors. Br J Psychiatry 2007; 190: 223–9.

25 Vinckier A, Saloppé X, Degouis F, Delaunoit B, Pham TH. Longitudinal follow-up of forensic patients: Focus on recidivism rate and risk scores. Ann Méd-Psychol Rev Psychiatr 2024; 182: 690–3.

26 Gibbon S, Huband N, Bujkiewicz S, Hollin CR, Clarke M, Davies S, et al. The influence of admission characteristics on outcome: Evidence from a medium secure forensic cohort. Personal Ment Health 2013; 7: 1–10.

27 Moher D, Liberati A, Tetzlaff J, Altman DG, The PRISMA Group. Preferred Reporting Items for Systematic Reviews and Meta-Analyses: The PRISMA Statement. PLOS Med 2009; 6: 336–41.

28 Stroup DF, Berlin JA, Morton SC, Olkin I, Williamson GD, Rennie D, et al. Meta-analysis of observational studies in epidemiology: A proposal for reporting. JAMA 2000; 283: 2008–12.

29 United Nations Office on Drugs and Crime. International Classification of Crime for Statistical Purposes, Version 1.0. United Nations Office on Drugs and Crime, 2015.

30 Munn Z, Moola S, Lisy K, Riitano D, Tufanaru C. Methodological guidance for systematic reviews of observational epidemiological studies reporting prevalence and cumulative incidence data. JBI Evid Implement 2015; 13: 147–53.

31 Andrade C. Need for and practical interpretations of the person-year construct in neuropsychiatric research. Indian J Psychol Med 2019; 41: 600–1.

32 Guevara JP, Berlin JA, Wolf FM. Meta-analytic methods for pooling rates when follow-up duration varies: a case study. BMC Med Res Methodol 2004; 4: 1–17.

33 Sterne J, Bodalia P, Bryden P, Davies P, López-López J, Okoli G, et al. Oral anticoagulants for primary prevention, treatment and secondary prevention of venous thromboembolic disease, and for prevention of stroke in atrial fibrillation: Systematic review, network meta-analysis and cost-effectiveness analysis. Health Technol Assess 2017; 21: 1–438.

34 Clark L, Zolotor A, Curteis T. MSR154 comparison of methods to estimate total person-time at risk for synthesis studies of incidence rates in observational data. Value Health 2024; 27: S468.

35 Higgins JPT, Thompson SG, Spiegelhalter DJ. A re-evaluation of random-effects meta-analysis. J R Stat Soc Ser A Stat Soc 2009; 172: 137–59.

36 Borenstein M. How to understand and report heterogeneity in a meta-analysis: The difference between I-squared and prediction intervals. Integr Med Res 2023; 12: 1–8.

37 Hunter JP, Saratzis A, Sutton AJ, Boucher RH, Sayers RD, Bown MJ. In meta-analyses of proportion studies, funnel plots were found to be an inaccurate method of assessing publication bias. J Clin Epidemiol 2014; 67: 897–903.

38 Viechtbauer W. Conducting meta-analyses in R with the metafor Package. J Stat Softw 2010; 36: 1–48.

39 39 Higgins JPT, Thomas J, Chandler J, Cumpston M, Li T, Page MJ, et al. Cochrane handbook for systematic reviews of interventions version 6.5. 2024. (www.cochrane.org/handbook).

40 Silver S, Cohen M, Spodak M. Follow-up after release of insanity acquittees, mentally disordered offenders, and convicted felons. Bull Am Acad Psychiatry Law 1989; 17: 387–400.

41 Russo G. Follow-up of 91 mentally ill criminals discharged from the maximum security hospital in Barcelona P.G. Int J Law Psychiatry 1994; 17: 279–301.

42 Baxter R, Rabe-hesketh, Sophia, Parroxtt J. Characteristics, needs and reoffending in a group of patients with schizophrenia formerly treated in medium security. J Forensic Psychiatry 1999; 10: 69–83.

43 Edwards J, Steed, Penny, and Murray K. Clinical and forensic outcome 2 years and 5 years after admission to a medium secure unit. J Forensic Psychiatry 2002; 13: 68–87.

44 Dowsett J. Measurement of risk by a community forensic mental health team. Psychiatr Bull 2005; 29: 9–12.

45 Simpson AIF, Jones RM, Evans C, McKenna B. Outcome of patients rehabilitated through a New Zealand forensic psychiatry service: a 7.5 year retrospective study. Behav Sci Law 2006; 24: 833–43.

46 Fioritti A, Melega V, Ferriani E, Rucci P, Venco C, Anna Rosa Scaramelli, et al. I percorsi assistenziali del paziente reo: il punto di osservazione dell’ospedale psichiatrico giudiziario. Noos 2006; 12: 91–5.

47 Simpson AIF, Chatterjee S, Duchcherer M, Ray I, Prosser A, Penney SR. Short-term outcomes for forensic patients receiving an absolute discharge under the Canadian Criminal Code. J Forensic Psychiatry Psychol 2018; 29: 867–81.

48 Probst T, Bezzel A, Hochstadt M, Pieh C, Mache W. Criminal recidivism after forensic psychiatric treatment. A multicenter study on the role of pretreatment, treatment-related, and follow-up variables. J Forensic Sci 2020; 65: 1221–4.

49 Marshall DJ, Vitacco MJ, Read JB, Harway M. Predicting Voluntary and Involuntary Readmissions to Forensic Hospitals by Insanity Acquittees in Maryland. Behav Sci Law 2014; 32: 627–40.

50 Davison S, Jamieson E, Taylor PJ. Route of discharge for special (high-security) hospital patients with personality disorder: Relationship with re-conviction. Br J Psychiatry 1999; 175: 224–7.

51 Friendship C, McClintock T, Rutter S, Maden A. Re-offending: patients discharged from a Regional Secure Unit. Crim Behav Ment Health 1999; 9: 226–36.

52 Dolan M, Khawaja A. The HCR–20 and post-discharge outcome in male patients discharged from medium security in the UK. Aggress Behav 2004; 30: 469–83.

53 Nicholls TL. Violence risk assessments with female NCRMD acquittees: Validity of the HCR-20 and PCL-SV.Dessertation Abstracts International: Section B: The Sciences and Engindeering 2004; 64: 4055.

54 Seifert DM-M S. Aktuelle Rückfalldaten der Essener prospektiven Prognosestudie [Preliminary recidivism rates of the Essener prognosis study]. Werd Deliktrückfälle Forensischer Patienten § 63 StGB Seltener 2005; 73: 16–22.

55 Philipse MWG, Koeter MWJ, van der Staak CPF, van den Brink W. Static and Dynamic patient characteristics as predictors of criminal recidivism: A prospective study in a dutch forensic psychiatric sample. Law Hum Behav 2006; 30: 309–27.

56 Yoshikawa K, Taylor PJ, Yamagami A, Okada T, Ando K, Taruya T, et al. Violent recidivism among mentally disordered offenders in Japan. Crim Behav Ment Health 2007; 17: 137–51.

57 Blattner R, Dolan M. Outcome of high security patients admitted to a medium secure unit: the Edenfield Centre study. Med Sci Law 2009; 49: 247–56.

58 Nilsson T, Wallinius M, Gustavson C, Anckarsäter H, Kerekes N. Violent recidivism: A long-time follow-up study of mentally disordered offenders. PLOS ONE 2011; 6: 1–9.

59 Bengtson S, Lund J, Ibsen M, Långström N. Long-term violent reoffending following forensic psychiatric treatment: Comparing forensic psychiatric examinees and general offender controls. Front Psychiatry 2019; 10: 1–11.

60 de Vogel V, Bruggeman M, Lancel M. Gender-sensitive violence risk assessment: Predictive validity of six tools in female forensic psychiatric patients. Crim Justice Behav 2019; 46: 528–49.

61 Nagata T, Tachimori H, Nishinaka H, Takeda K, Matsuda T, Hirabayashi N. Mentally disordered offenders discharged from designated hospital facilities under the medical treatment and supervision act in Japan: Reoffending and readmission. Crim Behav Ment Health 2019; 29: 157–67.

62 Haroon H, Wolfe N, Feizi S, Barboriak P. Assessing Two Decades of Insanity Acquittee Release from the North Carolina Forensic Program. J Am Acad Psychiatry Law Online 2023; 51: 1–11.

63 Ojansuu I, Latvala A, Kautiainen H, Forsman J, Tiihonen J, Lähteenvuo M. General and violent recidivism of former forensic psychiatric patients in Finland. Front Psychiatry 2023; 14: 1–6.

64 Sivak L, Forsman J, Masterman T. Duration of forensic psychiatric care and subsequent criminal recidivism in individuals sentenced in Sweden between 2009 and 2019. Front Psychiatry 2023; 14: 1–14.

65 Thomson L, Rees C. Long-term outcomes of the recovery approach in a high-security mental health setting: a 20-year follow-up study. Front Psychiatry 2023; 14: 1–19.

66 Bailey J, Macculloch M. Patterns of reconviction in patients discharged directly to the community from a special hospital: Implications for aftercare. J Forensic Psychiatry 1992; 3: 445–61.

67 Cope R, Ward M. What happens to Special Hospital patients admitted to medium security? J Forensic Psychiatry 1993; 4: 13–24.

68 Falla S, Sugarman P, Roberts L. Reconviction after Discharge from a Regional Secure Unit. Med Sci Law 2000; 40: 156–7.

69 Salem L, Crocker AG, Charette Y, Seto MC, Nicholls TL, Côté G. Supportive housing and forensic patient outcomes. Law Hum Behav 2015; 39: 311–20.

70 Argo D, Daibas K, Barash I, Abramowitz MZ. A 10-year comparison of short versus long-term court-ordered psychiatric hospitalization: A follow-up study. Isr J Health Policy Res 2023; 12: 1–7.

71 Hill C, Bagshaw R, Hewlett P, Perham N, Davies J, Maden A, et al. Estimating the effects of secure services on reconviction. part 1 – predictive validity of the offending groups reconviction scale (OGRS-2) and redundancy of patient social and clinical features. Int J Forensic Ment Health 2024; 23: 85–91.

72 Jewell A, Cocks C, Cullen AE, Fahy T, Dean K. Predicting time to recall in patients conditionally released from a secure forensic hospital: A survival analysis. Eur Psychiatry 2018; 49: 1–8.

73 Lyons G, Singh, Sara, Johnson, Anina, Nielssen, Olav, and Dean K. Rates and predictors of breach, revocation and hospital readmission of conditionally released forensic patients in New South Wales, Australia. J Forensic Psychiatry Psychol 2022; 33: 491–507.

74 Rossetto I, Clerici M, Franconi F, Felthous AR, Carabellese F, Di Vella G, et al. Differences between readmitted and non-readmitted women in an Italian forensic unit: A retrospective study. Front Psychol 2021; 12: 1–16.

75 Tellefsen C, Cohen MI, Silver SB, Dougherty C. Predicting success on conditional release for insanity acquittees: regionalized versus nonregionalized hospital patients. Bull Am Acad Psychiatry Law 1992; 20: 87–100.

76 Ojansuu I, Putkonen H, Lähteenvuo M, Tiihonen J. Substance abuse and excessive mortality among forensic psychiatric patients: A Finnish nationwide cohort study. Front Psychiatry 2019; 10: 1–17.

77 Takeda K, Sugawara N, Matsuda T, Shimada A, Nagata T, Kashiwagi H, et al. Mortality and suicide rates in patients discharged from forensic psychiatric wards in Japan. Compr Psychiatry 2019; 95: 1–8.

78 Pisani A. Long-term re-offending rates of adults and young people in NSW Bureau of Crime Statistics and Research, 2022 (https://bocsar.nsw.gov.au/documents/publications/bb/bb151-200/bb162-report-long-term-re-offending-rates-of-adults-and-young-people-in-NSW.pdf).

79 Gosek P, Kotowska J, Rowiñska-Garbieñ E, Bartczak D, Tomlin J, Heitzman J. Longer than prison? A comparison of length of stay in a medium security hospital and prison for perpetrators of violent crimes other than homicide or attempted homicide. Crim Behav Ment Health 2021; 31: 162–70.

80 Goodley G, Pearson D, Morris P. Predictors of recidivism following release from custody: A meta-analysis. Psychol Crime Law 2022; 28: 703–29.

81 Chi MH, Hsiao CY, Chen KC, Lee L-T, Tsai HC, Hui Lee I, et al. The readmission rate and medical cost of patients with schizophrenia after first hospitalization — A 10-year follow-up population-based study. Schizophr Res 2016; 170: 184–90.

82 Cook JA, Burke-Miller JK, Razzano LA, Steigman PJ, Jonikas JA, Santos A. Serious mental illness, other mental health disorders, and outpatient health care as predictors of 30-day readmissions following medical hospitalization. Gen Hosp Psychiatry 2021; 70: 10–7.

83 Zlodre J, Fazel S. All-cause and external mortality in released prisoners: Systematic review and meta-analysis. Am J Public Health 2012; 102: 67–75.

84 Tam J, Warner KE, Meza R. Smoking and the reduced life expectancy of individuals with serious mental illness. Am J Prev Med 2016; 51: 958–66.

85 Petersen SM, Toftdahl NG, Nordentoft M, Hjorthøj C. Schizophrenia is associated with increased risk of subsequent substance abuse diagnosis: A nation-wide population-based register study. Addiction 2019; 114: 2217–26.

86 Afzal M, Siddiqi N, Ahmad B, Afsheen N, Aslam F, Ali A, et al. Prevalence of overweight and obesity in people with severe mental illness: Systematic review and meta-analysis. Front Endocrinol 2021; 12: 1–12.

87 Aune D, Sen A, Norat T, Janszky I, Romundstad P, Tonstad S, et al. Body mass index, abdominal fatness, and heart failure incidence and mortality. Circulation 2016; 133: 639–49.

88 Petrelli F, Cortellini A, Indini A, Tomasello G, Ghidini M, Nigro O, et al. Association of obesity with survival outcomes in patients with cancer: A systematic review and meta-analysis. JAMA Netw Open 2021; 4: 1–30.

89 Campforts B, Drukker M, Crins J, van Amelsvoort T, Bak M. Association between antipsychotic medication and clinically relevant weight change: Meta-analysis. BJPsych Open 2023; 9: 1–11.

90 Martins LB, Monteze NM, Calarge C, Ferreira AVM, Teixeira AL. Pathways linking obesity to neuropsychiatric disorders. Nutrition 2019; 66: 16–21.

91 Huthwaite M, Elmslie J, Every-Palmer S, Grant E, Romans SE. Obesity in a forensic and rehabilitation psychiatric service: A missed opportunity? J Forensic Pract 2017; 19: 269–77.

92 Samuels E, Moran N. Accessing and engaging with primary health care services following discharge from forensic secure services: The perspectives of service users and mental health practitioners. J Forensic Pract 2021; 23: 117–31.

93 Chung DT, Ryan CJ, Hadzi-Pavlovic D, Singh SP, Stanton C, Large MM. Suicide rates after discharge from psychiatric facilities: A systematic review and meta-analysis. JAMA Psychiatry 2017; 74: 694–702.

94 Swaraj S, Wang M, Chung D, Curtis J, Firth J, Ramanuj PP, et al. Meta-analysis of natural, unnatural and cause-specific mortality rates following discharge from in-patient psychiatric facilities. Acta Psychiatr Scand 2019; 140: 244–64.

95 Lawrence D, Bagshaw R, Stubbings D, Watt A. Restrictive practices in adult secure mental health services: A scoping review. Int J Forensic Ment Health 2022; 21: 68–88.

96 Rücker G, Schwarzer G, Carpenter JR, Binder H, Schumacher M. Treatment-effect estimates adjusted for small-study effects via a limit meta-analysis. Biostatistics 2011; 12: 122–42.

