## Supplementary Analyses for "Life Beyond the Forensic Unit: A Systematic Review and Meta-analysis of Patient Reoffending, Hospital Readmission, and Mortality Rates Following Discharge to the Community"

### Appendix A: Sensitivity analysis

A post-hoc random-effect size meta-analysis examining violent reoffending incidence was performed to examine rates after 2013. Eleven studies reported on violent reoffending among patients discharged from secure care [2,4,22,47,59–65]. A random-effects model showed that crude violent reoffending rates range from 145 to 16,667 per 100,000 person-years, with a pooled estimate of 1,416, 95% CI [680, 2,949], $I^2$ = 94.3%, 95% CI [91.6% - 96.2%]. In terms of heterogeneity, the 95% prediction interval ranged from 85 to 23,436.

A separate post-hoc random-effect size meta-analysis assessing reconviction incidence was performed to examine rates after 2013. A total of twelve studies reported on reconvictions among discharged forensic patients [1,2,4,59–65,69]. A random-effects model showed that crude violent reoffending rates range from 614 to 7,638 per 100,000 person-years, with a pooled estimate of 2,612 95% CI [1757, 3881], $I^2$ = 97.4%, 95% CI [96.5% - 98.1%]. In terms of heterogeneity, the 95% prediction interval ranged from 548 to 12,440.

An additional random-effect size meta-analysis examining readmission incidence was performed to examine rates after 2013. Fifteen studies reported on readmission rates among patients discharged from secure care [1,16,21,23,47,49,60–62,69–74]. A random-effects model showed that crude readmission rates range from 977 to 27,586 per 100,000 person-years, with a pooled estimate of 8314, 95% CI [5,047, 13,697], $I^2$ = 99.3%, 95% CI [99.2% - 99.4%]. In terms of heterogeneity, the 95% prediction interval ranged from 960 to 72,018.
